# The kinetic variations of anti-nucleocapsid antibody in SARS-CoV-2 infection

**DOI:** 10.1101/2021.02.05.21251208

**Authors:** Shoji Kawada, Atsushi Ogata, Yasuhiro Kato, Masashi Okamoto, Yuta Yamaguchi, Takayoshi Morita, Atsushi Kumanogoh

## Abstract

The humoral immune response to severe acute respiratory syndrome coronavirus 2 (SARS-CoV-2) plays a pivotal role in controlling coronavirus disease 2019 (COVID-19) infections. However, little is known about the persistence of the antibody response.

We evaluated that the kinetics of anti-nucleocapsid protein antibody of SARS-CoV2 infected healthcare workers in COVID-19 cluster occurred hospital. The long-term kinetics of anti-N antibody was classified high and keep pattern, high and decay pattern, and low and keep pattern. COVID-19 contact and symptomaticity was not related to kinetic patterns.

The reason of kinetic difference was still unclear. However natural anti-SARS-CoV-2 antibody persistence was not uniform, suggesting inter-individual difference of SARS-CoV2 vaccine efficacy.

## Introduction

The humoral immune response to severe acute respiratory syndrome coronavirus 2 (SARS-CoV-2) plays a pivotal role in controlling coronavirus disease 2019 (COVID-19) infections. Serologic tests that measure antibodies are the preferred method for evaluating immunity to SARS-CoV-2. However, little is known about the persistence of the antibody response.

## Methods

A local outbreak of COVID-19 occurred between April 11 to May 11 at the Diani-Osaka Police Hospital in Japan. During the outbreak, comprehensive cross-sectional nucleic acid tests were performed in health care workers (HCWs) from April 27 to May 11. Subsequent serologic surveys were performed using a semi-quotative assay for anti-nucleocapsid (N) immunoglobulin antibodies (Roche Diagnostics) from May 25 to June 16. Monthly follow-up serologic surveys were conducted in seropositive individuals. Awareness of contact with COVID-19 and related symptoms were measured using questionnaires. Anti-spike (S) antibodies (Roche) were evaluated 7 months after diagnosis.

## Results

Of the 576 hospital employees, 520 HCWs underwent nucleic acid tests and serologic tests for SARS-CoV-2. 42 HCWs were nucleic acid tested positive. During the outbreak at the hospital, the first serologic tests were performed 22.2 ± 8.4 days after the nucleic acid test, of which 52 were seropositive. Monthly follow-up serologic surveys were performed in 45 seropositive HCWs. The kinetics of anti-N antibodies were classified into three patterns (Figure 1). In 15 individuals, anti-N antibodies significantly increased and persisted (high-keep); in 16 individuals, anti-N antibodies increased significantly but later declined (high-decay); and in 14 individuals, anti-N antibodies increased modestly and persisted (low-keep). Furthermore, anti-N antibodies increased gradually over time in six individuals (three with the high-keep pattern and three with the low-keep pattern). Individuals with the high-keep kinetic pattern of anti-N antibodies had a higher titer of anti-S antibodies at 7 months postdiagnosis. The kinetic pattern of anti-N antibodies showed no apparent correlation with either COVID-19 contact or symptomaticity (Table 1). Seropositivity persisted in all HCWs up to 7 months after diagnosis, and there were no reinfections of COVID-19.

**Figure-1.**
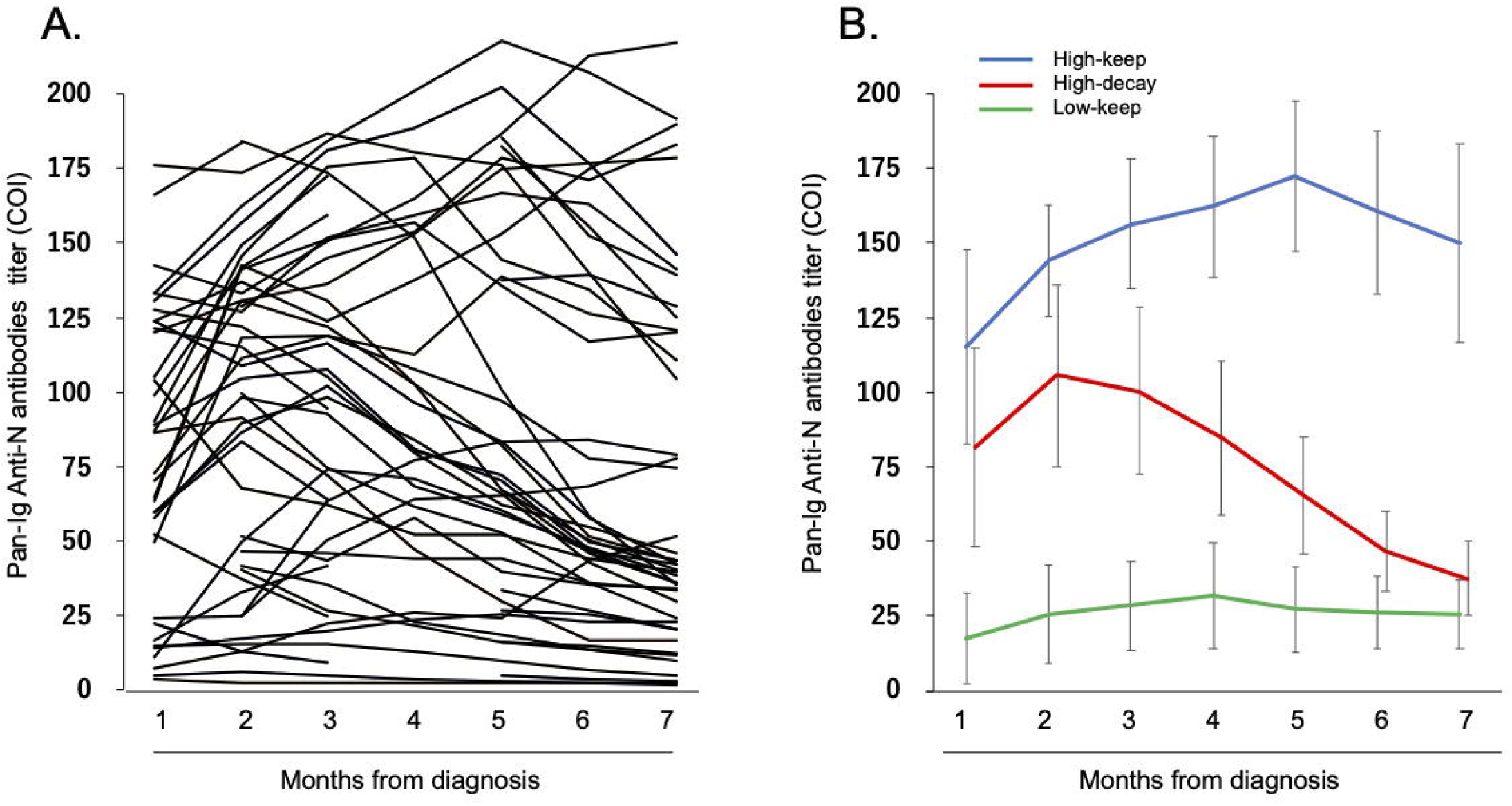
kinetics patterns of antibody against to SARS-CoV-2 N protein.

## Discussion

Most SARS-CoV-2 infected individuals display an antibody response within 21 days of infection. However, the persistence of antibodies against SARS-CoV-2 remains controversial. Some studies have reported robust antibody responses against SASR-CoV-2 (1,2), whereas others have reported declines in antibody titers (3, 4). In our study, longitudinal change in anti-N antibodies was not uniform and was classified into three patterns: high levels of anti-N antibodies were obtained and maintained (high-keep); high levels of anti-N antibodies were obtained but declined (high-decay); and low levels of anti-N antibodies were obtained and maintained (low-keep). Antibody titers increased in some individuals. In our cohort, all HCWs were exposed to the COVID-19 outbreak at the hospital simultaneously. Additional close contact to COVID-19 was not reported in any HCWs. The antibody response was not boosted by re-exposure to SARS-CoV-2. The cause of kinetic variation of the anti-N antibody response was unclear but may be influenced by unknown interindividual differences in antibody response.

Previous reports have demonstrated that severe infections exhibit a strong and prolonged antibody response (5). Anti-S antibodies were enriched among recovering individuals, whereas anti-N antibodies were enriched in deceased individuals (6). In our study, disease severity was not correlated with the titer or kinetic pattern of anti-N antibodies. Because the nucleocapsid protein of the COVID-19 virus is not exposed to the outside the virion, anti-N antibodies respond to destroyed or incomplete virus particles but not to infectable virus particles. Therefore, anti-N antibodies may not be related to disease recovery. The variation in antibody response raises questions concerning the efficacy of the SARS-CoV-2 vaccine. A personalized approach to boost the immune response to the SARS-CoV-2 vaccine may be necessary to eradicate COVID-19.

## Supporting information

COI(Kato)

COI(Kawada)

COI(Kumanogoh)

COI(Morita)

COI(Ogata)

COI(Okamoto)

COI(Yamaguchi)

## Data Availability

The data that support the findings of this study are available from the corresponding author upon reasonable request.

## Acknowledgments

The authors would like to thank Enago (www.enago.jp) for the English language review.

---

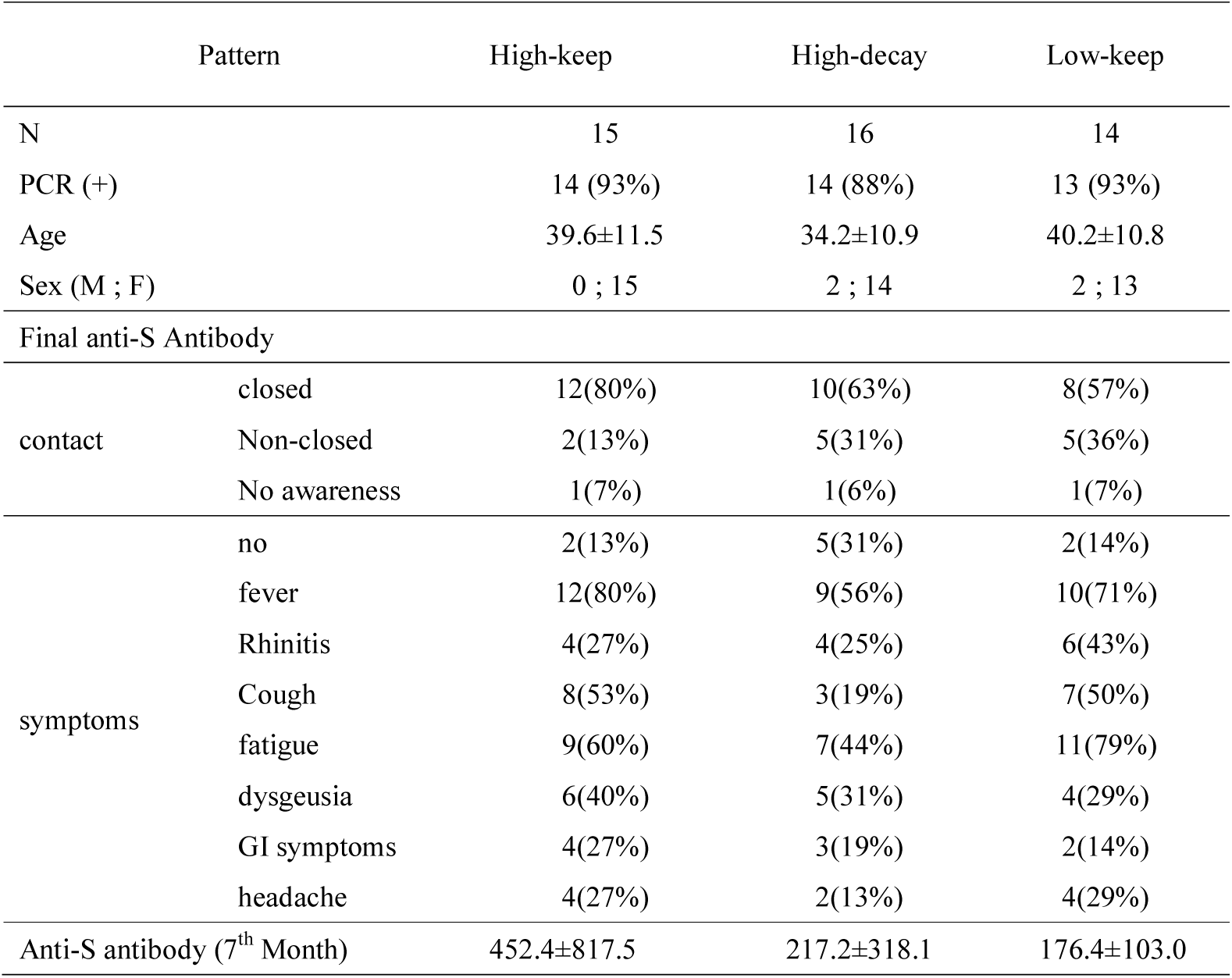

